# Linkage to primary care public health facilities for cardiovascular disease prevention : A community based cohort study from urban slums in India

**DOI:** 10.1101/2020.09.21.20198622

**Authors:** Abhijit P Pakhare, Ankur Joshi, Rasha Anwar, Khushbu Dubey, Sanjeev Kumar, Shubham Atal, Ishan Raj Tiwari, Vipul Mayank, Neelesh Shrivastava, Rajnish Joshi

**Author notes:** Author Email Corresponding Author - Dr Rajnish Joshi, Department of Medicine, All India Institute of Medical Sciences, Bhopal.

## Abstract

**Objectives:** Hypertension and diabetes mellitus are key risk factors for Cardiovascular diseases. Pharmacotherapy and life-style modifications are necessary. Once screened, individuals need to be linked to primary health-care system for initiation and maintenance of therapies, to achieve optimal blood pressure and glycemic control. In the current study we evaluate predictors and barriers for non-linkage to primary-care public health facilities for CVD risk reduction.

**Methods:** We conducted a community-based longitudinal study in 16 urban slum clusters in central India. Community health workers (CHWs) in each urban slum cluster screened all adults aged 30 years or more for hypertension and diabetes, and those positively screened were sought to be linked to Urban Primary Health Centres (UPHCs). We performed univariate and multivariate analysis to identify independent predictors for non-linkage to primary-care providers. We conducted in-depth assessment in 10% of all positively screened, to identify key barriers that potentially prevented linkages to primary-care facilities.

**Results:** Of 6174 individuals screened 1451(23.5%; 95%-CI 22.5-24.6) were identified as high-risk, and required linkage to primary-care facilities for pharmacotherapy. Out of these, 544(37.5%) were linked to public primary-care facilities, 259(17.9%) to private providers, 142(9.8%) were treatment interrupters, and 506(34.9%) didn’t get linked to any provider. On multivariate analysis, as compared to those linked to public primary care facilities, those who were not linked had age less than 45 years (OR 2.2 (95%CI 1.3-3.5)); were in lowest wealth quintile (OR 1.8 (95%CI 1.1-2.9); resided beyond a kilometre from UPHC (OR 1.7 (95%CI 1.2-2.4); and were engaged late by CHWs (OR 2.6 95%CI (1.8-3.7)). Despite having comparable knowledge level, denial about their risk-status and lack of family support were key barriers in this group.

**Conclusions:** This study highlights importance of early engagement through CHWs after positive screening, strategies to engage with younger individuals who may be in denial about their risk-status.

**Article summary - Strengths and limitations of this study:**

- This is community based longitudinal study implemented through community health workers (CHWs).
- It is “real-world” implementation as per national non-communicable disease control program in India (known as NPCDCS), which envisages population based screening through community health workers (CHWs), and linkages to public health facilities.
- This study highlights that within urban slum, being young, in a low socioeconomic position, distance from health facility are important determinants of linkage to public health facility.
- Early engagement by CHWs enhances likelihood of linkage.
- This study was limited to urban slum clusters from a single city, however we believe that health-infrastructure is broadly similar in such settings elsewhere.

## Introduction

Hypertension, and diabetes mellitus are important risk factors for cardiovascular disease (CVD), and achieving optimal blood-pressure and glycemic control is challenging.[1] In urban India about 30-40% of all adults have hypertension and about 10-15% have diabetes mellitus.[2,3] Only half of the individuals with hypertension are aware of their elevated blood pressures, and of those aware, half are not on any treatment. Further, only about half of all individuals with hypertension who are on medication are controlled.[4] In order to bridge awareness-treatment-control gaps, Indian National Program for Control of Diabetes, Cancer and Stroke (NPCDCS) was initiated in year 2010. This program envisages to annually screen all adults aged 30 years and above for presence of hypertension and diabetes mellitus, initiate life-style changes and drug therapy in those positively screened, and follow them up for treatment adherence.[5] Availability and affordability of preventive drug therapies was a key barrier in achieving control of risk factors. [6] In order to overcome this barrier, NPCDCS has made medications for blood-pressure and glycemic control available in public sector primary care facilities. Various other evidence-practice gaps however remain a challenge for CVD prevention. [7]

CVD prevention requires a multi-level approach. Individuals who are identified with hypertension and diabetes mellitus in the community are largely asymptomatic.[8,9] At individual level there is a need to adopt various life-style measures to reduce CVD-risk, which requires sustained behaviour change. Cultural acceptance of behavioural change is required at a societal level especially for risk factors such as tobacco use, physical inactivity and dietary practices. Drug therapy for blood-pressure and glycemic control is an integral part of CVD prevention, and making these available requires health-system-level strengthening. Despite improving availability and affordability of drug therapy, individual-level (related to knowledge, attitude, beliefs and practices) and health-system-level barriers (infrastructure, access, and quality of services) remain important.[10] High-income countries have a much better availability and affordability of anti-hypertensive medication, yet their blood-pressure control is only marginally better (36% in high income vs 23% in low-income countries).[6] Individuals with hypertension and diabetes need to be effectively linked to primary-care facilities so that their blood-pressures and sugars are controlled, and their CVD risk is lowered. For this linkage to be successful, individual, societal, and health system level barriers need to be addressed.

Linkage of individuals identified with hypertension and diabetes to primary health-care facilities is an important step towards CVD risk reduction. It is important to understand which high risk individuals do not get linked and predictors of non-linkage. We have explored these issues in the current study, where a community-wide screening was performed as envisaged in NPCDCS, and high-risk individuals were sought to be linked to public sector primary care facilities.

## Methods

### Design and Ethics Statement

We designed a community based longitudinal study to identify predictors of non-linkage to public-health primary care facilities. The study design was approved by the institutional human ethics committee (Ref: IHEC-LOP/2017/EF00045) and funded by Indian Council of Medical Research. All participants provided written informed consent prior to initiation of any study procedures. Detailed study protocol is available on pre-print server.[11]

### Setting

The study was conducted in 16 urban slum clusters from catchment area of two Urban Primary Health Centres (UPHCs) of Bhopal, a city located in central India. These UPHCs were located at *Barkheda Pathani and Saibaba Nagar* which are usual places from where study participants sought primary healthcare. Every urban slum cluster has a designated accredited social health activist (ASHA) who functions as a community health worker (CHW), and provides linkages to the public health delivery system. In addition to UPHCs, primary care needs in public sector are also met by the district hospital, and government owned medical college hospitals. There are no out-of-pocket costs towards either consultation or available medications at these facilities. Many individuals opt to seek care from private sector, which is larger both in terms of number of providers, as well as individuals who seek care through it. Every primary-care consultation in private sector incurs out-of-pocket expenditure (range 1 to 10 USD), and prescribed medications are available through private pharmacies. Private care providers have longer working hours, and could be located in closer proximity to some urban slum clusters.

### Participants

All individuals above 30 years of age and residing in the selected clusters for 6 months or more were eligible for inclusion. Women who were pregnant at the time of screening were excluded. No other exclusions were applied. We identified individuals with hypertension (previously known hypertension or SBP > 140 or DBP >90 mm Hg on two or more occasions), diabetes (Previously known diabetes or Random blood sugar > 200mg/dL or a fasting blood sugar > 126mg/dL), a previous known cardiovascular or a cerebrovascular event, and classified them as high-CVD risk. This high-risk group was followed up over next eighteen months to demonstrate feasibility of CHW based interventions.

### Study Procedures

Detailed study procedures are described elsewhere.[11] Briefly these included the following:

### CHW recruitment and training

ASHAs who function as CHW in each cluster were trained in basic health issues with respect to CVD, its risk factors, and their prevention. The training consisted of interview and communication techniques, measuring physical parameters like height, weight, waist circumference (WC), blood pressure measurement using a digital sphygmomanometer, and blood sugar estimation using a glucometer. After training of CHWs we organized community mobilization meetings to sensitize people regarding roll out of the study. These community mobilization meetings were organized by CHW of the concerned area, and Medical Social Worker (Project Supervisor) and were attended by study investigator/s. These meetings improved cooperation from the communities.

### Screening for CVD risk factors

CHWs administered a screening consisted of a questionnaire (to identify tobacco or alcohol consumption, physical activity levels, previously known hypertension, diabetes or a manifest CVD such as ischemic heart disease or stroke), anthropometry (to measure body mass index (BMI) and waist circumference (WC) (weighing scale-Seca-876, stadiometer-Seca-213 and measuring tapes Seca-201, Seca, Hamburg, Germany), measurement of blood-pressure (using Omron digital apparatus model 7200, Kyoto, Japan) and a non-fasting blood glucose (RBG) by glucometer (SD diagnostics, Korea). Within one week of a set of screenings performed by CHWs through home-visits, a diagnosis confirmation camp was conducted by the study supervisor to obtain a second set of blood pressure readings and a random blood glucose assessment. Risk stratification decision was made based on standard operational definitions of elevated blood pressure and blood sugar levels by the study physicians. To ensure quality-check, supervisors used to cross-check 10% of blood pressure measurements, tobacco and previously known NCD questions. All data was collected on mobile phone based tool-Commcare (Dimagi Inc, USA). All CHWs were provided with an android based mobile phone with installed Commcare application for data collection. This tool has a facility to track visits and details of the previous visits can be made available. We developed a decision support system for identification of individuals at high risk for future CVD as per the operational definitions (see below). Questionnaire used in the study are available in supplementary file.

### Management of “high-risk” individuals

All “high risk” individuals were advised about tobacco cessation, dietary modification, and increase in physical activity by CHWs and study supervisors. While all individuals were free to seek care from either the nearest UPHC or any other source of their choice, they were provided a referral-slip with previous blood pressure and blood sugar values to facilitate subsequent decision making. If they chose to visit the UPHC, they were evaluated by a physician. UPHC is functional between 12 noon and 5pm on all weekdays, with one day in a week designated as non-communicable disease clinic (Tuesdays for *Saibaba nagar*, and Wednesday for *Barkheda Pathani* UPHCs). The physicians at UPHC were trained to follow simple therapeutic algorithms for treatment initiation, optimization and continuation for hypertension (Based on JNC 8 hypertension guidelines)[12], and diabetes (based on ADA guidelines 2017).[13] Blood pressure of all participants, and blood sugar levels of participants were measured at UPHC for reconfirmation and facilitation of treatment decisions. The physician advised either treatment initiation (if they were newly detected with hypertension or diabetes mellitus), treatment optimization (if they were previously known to have these risks but were not controlled (SBP >140; DBP >90, FBS >140; PPBS>180)) or treatment-continuation (if they were well controlled on previous therapies). The physicians could choose from available drugs at UPHC which were Losartan (Angiotensin receptor blocker), Amlodepine (calcium channel blocker), Hydrocholorthiazide (diuretic), Metformin (Biguanide), Glimiperide (sulphonylurea), low-dose Aspirin, and Atorvastatin. These drugs have also been identified by WHO Package of Essential Noncommunicable Disease Interventions (WHO PEN) as essential ingredients for NCD care.[14] Individuals were usually dispensed with 15 days of drug therapy, and advised for a refill thereafter. Those individuals who would not be optimally controlled on maximal permissible dosages of available drugs, despite adequate drug adherence were advised consultation with specialists at secondary or tertiary care hospitals in the public sector. All these treatment decisions (Initiation, escalation, de-escalation of drug therapy) were recorded by the study physician in a NCD register available at UPHC for the research project, which also was a visit-log for high risk participants. Individuals who were linked to UPHC were identified from this NCD register, which was maintained in hard copy. The data of the NCD register was updated weekly into a designated data collection software by study supervisors. Periodic data quality check was done by study investigators. Those who had three or more visits logged in six months from their initial visit at UPHC were classified as “treatment continuers”, and those who had fewer visits were “treatment-interrupters”.

### Follow-up visits by CHWs and outcome classification

Subsequent to initial screening, CHWs performed home-visits, once in every two months to reinforce linkages to public health facilities and adherence to drug-treatment. In the six-month home-visit (third visit), CHWs again recorded source of CVD-prevention treatment, and identified individuals who were “not-on treatment”, “on treatment from private care providers”, and “treatment-continuers” or “treatment-interrupters”. Based on this information and treatment records available from UPHC-NCD register all high-risk individuals were classified into four groups: group A, linked to public sector primary care facilities and treatment continuers; group B, linked to private care providers and treatment continuers; group C, linked to public or private sector primary care facilities and treatment interrupters; group D not-linked to any provider.

### Patient and public involvement

This study was implemented through Community Health Workers (ASHAs) who reside in the same community. After training of CHWs we have organized community mobilization meetings to sensitize people regarding roll out of the study. These community mobilization meetings were organized by CHW of the concerned area and Medical Social Worker (Project Supervisor) and were attended by study investigator/s. These meetings ensured cooperation of public for the conduct of study.

### Operational Definitions

Optimal blood pressure control was defined as SBP<140 and DBP<90 mm of Hg. Optimal glycemic control was defined as fasting blood sugar < 126 mg/dl and post-prandial blood sugar < 180 mg/dl or HbA1c < 7. “ASHA engagement” has been classified based on the home-visit to confirmation camp-visit interval. Confirmation visit happening within 7 days has been defined as “early engagers”, between one week and one month were “intermediate” and beyond one month were “late-engagers”. Outcome was classified in terms of the participants being linked to public health public health facilities, or linked to public health facilities but interrupted, or linked to private health facilities, or not linked to any facilities.

### Sampling for identification of barriers to linkage

Barrier identification was done after twelve months of follow up period. For exploring facilitators and barriers of linkage, a semi-structured questionnaire was prepared which has items categorized in different domains (Knowledge, Attitude, Individual, Health Provider and Health System). Respondents had to indicate whether a particular item is applicable to them or not. It also has open ended questions about reasons for non-linkage. This questionnaire was administered to sub-sample from each category of participants. For this, we performed a stratified random sampling and listed numerically proportionate individuals from each of the four groups A, B, C, and D, ensuring a sample size of 10 % from each group.

### Statistical Analysis

All data analysis was done using SPSS software (IBM SPSS Statistics for Macintosh, Version 26.0. Armonk, NY: IBM Corp.). Wealth index was constructed by using principal component analysis on household asset ownership data.[15] Households were classified in different wealth quintiles, higher the wealth quintile indicates relatively better socio-economic position. Distribution of continuous variables across linkage-groups was compared using ANOVA and dichotomous variables using chi-square test. A p-value of <0.05 was considered as significant for these comparisons. To determine predictors of non-linkage and linkage to private facility, individuals in group A (linked to public health facilities) were considered as reference. Considering more than two categories of outcome variable (Linkage Status) we used multi-nominal logistic regression model to identify variables independently associated with non-linkage, linkage to private facilities and interrupters after linkage with reference to linkage to public health facilities. Odds ratio (OR) was estimated to represent point-estimate and its 95% confidence interval as a measure of precision of the association. We performed a descriptive analysis of the barriers in each of the domains, and for individuals in each of the outcome categories.

## Results

Between November 2017 and June 2018 a total of 6174 individuals were screened, and 1451 participants (23.5%; 95%-CI 22.5-24.6) were identified as high-risk. Study flow is depicted in Figure 1. Most of these individuals were middle aged, and women (n=858, 59.2%). Six months after initial screening, 55.2% of all high-risk individuals were linked and were treatment-continuers. Of these 544 (37.5%) were linked to public health facilities (Group A), and 259 (17.9%) to private care providers (Group B). Another 142 (9.8%) were treatment-interrupters (Group C), and 506 (34.9%) never got linked to any provider (Group D). Most individuals in Group A (linked to public facilities) were of age more than 45 years (69.3 %), were women (67.8 %) and lived within 3km of UPHC (72.6%), did not use tobacco (60.1%). About half of them (44.9%) were engaged early by ASHAs. Characteristics of individuals in these four groups are presented in Table 1.

**Table 1:** Distribution of socio-demographic, risk factors, measurements and classification based on linkage status

|  |  | Group A | Group B | Group C | Group D | p-value |
| --- | --- | --- | --- | --- | --- | --- |
| Variables |  | Linked to Public Health Facility<br>N=544 | Linked to Private providers<br>N=259 | Treatment Interrupters<br>N=142 | Not linked<br>N=506 |  |
| Age Group | <= 45 | 167 (30.7) | 88 (34) | 57 (40.1) | 250 (49.4) | <0.001 |
|  | 46 - 65 | 286 (52.6) | 129 (49.8) | 72 (50.7) | 195 (38.5) |  |
|  | 66+ | 91 (16.7) | 42 (16.2) | 13 (9.2) | 61 (12.1) |  |
| Gender | Male | 175 (32.2) | 110 (42.5) | 61 (43) | 247 (48.8) | <0.001 |
|  | Female | 369 (67.8) | 149 (57.5) | 81 (57) | 259 (51.2) |  |
| Formal Education | Yes | 283 (52) | 167 (64.5) | 80 (56.3) | 311 (61.5) | 0.002 |
|  | No | 261 (48) | 92 (35.5) | 62 (43.7) | 195 (38.5) |  |
| Marital Status | Other | 121 (22.2) | 48 (18.5) | 28 (19.7) | 90 (17.8) | 0.308 |
|  | Married | 423 (77.8) | 211 (81.5) | 114 (80.3) | 416 (82.2) |  |
| Distance form PHC | More than 3 km | 149 (27.4) | 77 (29.7) | 50 (35.2) | 155 (30.6) | 0.001 |
|  | 1-3 km | 145 (26.7) | 67 (25.9) | 39 (27.5) | 181 (35.8) |  |
|  | < 1 km | 250 (46) | 115 (44.4) | 53 (37.3) | 170 (33.6) |  |
| Current Oral Tobacco Use | Yes | 217 (39.9) | 105 (40.5) | 65 (45.8) | 247 (48.8) | 0.020 |
|  | No | 327 (60.1) | 154 (59.5) | 77 (54.2) | 259 (51.2) |  |
| Current Smoking | Yes | 35 (6.4) | 22 (8.5) | 14 (9.9) | 54 (10.7) | 0.098 |
|  | No | 509 (93.6) | 237 (91.5) | 128 (90.1) | 452 (89.3) |  |
| Current Alcohol use | Yes | 69 (12.7) | 33 (12.7) | 28 (19.7) | 139 (27.5) | <0.001 |
|  | No | 475 (87.3) | 226 (87.3) | 114 (80.3) | 367 (72.5) |  |
| BMI<sup>#</sup> | >=25 | 285 (52.9) | 119 (56.1) | 58 (49.6) | 175 (48.3) | 0.283 |
|  | <25 | 254 (47.1) | 93 (43.9) | 59 (50.4) | 187 (51.7) |  |
| Abdominal Obesity | Yes | 383 (70.4) | 194 (74.9) | 87 (61.3) | 306 (60.5) | <0.001 |
|  | No | 161 (29.6) | 65 (25.1) | 55 (38.7) | 200 (39.5) |  |
| New HTN Diagnosis | Yes | 171 (31.4) | 59 (22.8) | 72 (50.7) | 207 (40.9) | <0.001 |
|  | No | 373 (68.6) | 200 (77.2) | 70 (49.3) | 299 (59.1) |  |
| New DM Diagnosis | Yes | 51 (9.4) | 14 (5.4) | 13 (9.2) | 56 (11.1) | 0.087 |
|  | No | 493 (90.6) | 245 (94.6) | 129 (90.8) | 450 (88.9) |  |
| Engagement by ASHA<sup>#</sup> (Home visit to camp interval) | Late (>1 month) | 116 (21.5) | 74 (34.9) | 32 (27.4) | 156 (43.1) | <0.001 |
|  | Intermediate | 181 (33.6) | 49 (23.1) | 40 (34.2) | 69 (19.1) |  |
|  | Early (<7 days) | 242 (44.9) | 89 (42) | 45 (38.5) | 137 (37.8) |  |
| Wealth Index Quintile<sup>##</sup> | Q1 | 67 (13.1) | 21 (8.4) | 23 (17.7) | 77 (16.6) | 0.001 |
|  | Q2 | 79 (15.4) | 32 (12.9) | 21 (16.2) | 86 (18.5) |  |
|  | Q3 | 101 (19.7) | 46 (18.5) | 27 (20.8) | 88 (18.9) |  |
|  | Q4 | 114 (22.3) | 49 (19.7) | 33 (25.4) | 101 (21.7) |  |
|  | Q5 | 151 (29.5) | 101 (40.6) | 26 (20) | 113 (24.3) |  |

All numbers indicate frequency (proportion) unless indicated otherwise. # Data is missing for 221 participants (5, 47, 25 and 144 respectively in Groups A, B, C and D).
Data is missing for 95 participants (32, 10, 12 and 41 respectively in Groups A, B, C and D).
Formal education- Any level of formal education in school; Tobacco use- use of any tobacco product in last 30 days ; Abdominal obesity- Waist circumference more than 90 cm for men and 80 cm for women.

**Figure 1.**
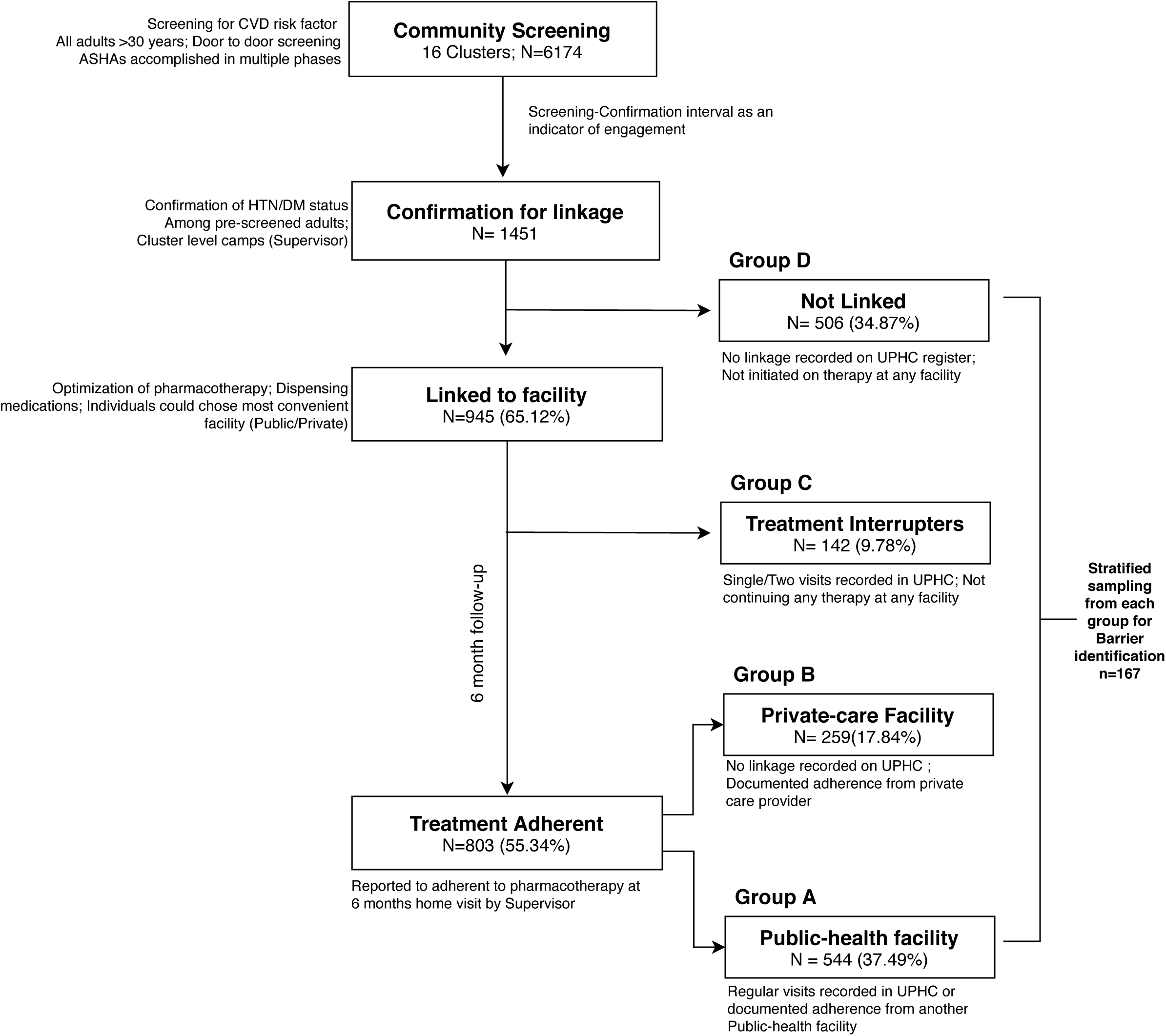
Study Flow. Abbreviations in figure: ASHA Accredited social health activist; UPHC Urban Primary Health Center; CVD Cardiovascular disease; HTN Hypertension; DM Diabetes mellitus;

As compared to those who were linked to public health facilities, those who never got linked (Group D) were more likely to be young (OR 2.17 (95%CI 1.33-3.53)), in lowest wealth quintile (OR 1.8 (95%CI 1.1-2.9)), consumed alcohol (OR 1.9 (95%CI 1.25-2.96)). These individuals also engaged late with CHWs (OR 2.6 (95%CI 1.84-3.7)), and lived farther away from UPHC (OR 1.7 (95%CI 1.19-2.42)). These risk estimates are derived from multi-nominal logistic regression analysis. Goodness of fit was statistically non-significant (p=0.906), likelihood ratio test for model fitting criteria was statistically significant (p<0.001; Cragg-Uhler(Nagelkerke) R2=0.623). This model fitting information indicates that current model can be used to understand predictors of linkage status. (Table 2)

**Table 2:** Unadjusted and adjusted multi-nominal regression for predictors of non-linkage

| Variable | Group B<br>Linked to Private OR (95%CI) |  | Group C<br>Treatment interrupters OR (95%CI) |  | Group D<br>Not linked OR (95%CI) |  |
| --- | --- | --- | --- | --- | --- | --- |
|  | Unadjusted | Adjusted | Unadjusted | Adjusted | Unadjusted | Adjusted |
| Age |  |  |  |  |  |  |
| More than 65 years (Elderly) | Ref |  | Ref |  | Ref |  |
| Between 30-45 years (Young) | 1.16 (0.85-1.59) | 1.16 (0.67-2) | 1.51 (1.03-2.22) | 2.34 (1.08-5.06) | 2.2 (1.71-2.84) | 2.17 (1.33-3.53) |
| Between 46-65 years (Middle aged) | 0.9 (0.67-1.2) | 1 (0.61-1.64) | 0.93 (0.64-1.34) | 1.74 (0.84-3.59) | 0.57 (0.44-0.72) | 1.1 (0.69-1.74) |
| Gender |  |  |  |  |  |  |
| Women |  |  |  |  |  |  |
| Men | 1.56 (1.15-2.11) | 1.63 (1.04-2.57) | 1.59 (1.09-2.32) | 1.38 (0.76-2.51) | 2.01 (1.57-2.58) | 1.45 (0.96-2.17) |
| Formal schooling (vs no formal schooling) | 1.67 (1.23-2.27) | 1.39 (0.93-2.06) | 1.19 (0.82-1.73) | 0.85 (0.51-1.4) | 1.47 (1.15-1.88) | 1.12 (0.79-1.59) |
| Economic status (Wealth Quintile) |  |  |  |  |  |  |
| First Quintile (Lowest) | 0.61 (0.37-1.02) | 0.6 (0.31-1.15) | 1.43 (0.85-2.4) | 2.32 (1.09-4.93) | 1.32 (0.92-1.88) | 1.8 (1.1-2.96) |
| Second Quintile | 0.81 (0.52-1.26) | 0.76 (0.44-1.3) | 1.06 (0.62-1.79) | 1.66 (0.77-3.59) | 1.24 (0.89-1.74) | 1.57 (0.99-2.49) |
| Third Quintile | 0.92 (0.63-1.36) | 0.87 (0.54-1.4) | 1.07 (0.66-1.72) | 2.02 (1.02-4.01) | 0.95 (0.69-1.31) | 0.97 (0.62-1.52) |
| Fourth Quintile | 0.86 (0.59-1.25) | 0.71 (0.45-1.13) | 1.19 (0.76-1.86) | 2.19 (1.12-4.28) | 0.97 (0.72-1.31) | 1.08 (0.7-1.66) |
| Fifth Quintile (Highest) |  |  |  |  |  |  |
| Current Oral Tobacco use (vs Non-use) | 1.03 (0.76-1.39) | 1.04 (0.7-1.54) | 1.27 (0.88-1.85) | 0.76 (0.46-1.27) | 1.44 (1.13-1.84) | 1 (0.71-1.41) |
| Current Smoking (vs Non-smoking) | 1.35 (0.77-2.35) | 1.15 (0.57-2.31) | 1.59 (0.83-3.05) | 2.06 (0.91-4.66) | 1.74 (1.11-2.71) | 1.15 (0.64-2.06) |
| Current Alcohol use (vs no use) | 1.01 (0.64-1.57) | 0.72 (0.41-1.26) | 1.69 (1.04-2.74) | 0.77 (0.38-1.55) | 2.61 (1.89-3.59) | 1.92 (1.25-2.96) |
| Engagement by ASHA (Home visit to camp interval) |  |  |  |  |  |  |
| Early (< 7 days) |  |  |  |  |  |  |
| Intermediate (7 days to 1 month) | 0.59 (0.41-0.86) | 0.68 (0.45-1.04) | 1.03 (0.67-1.57) | 1.21 (0.73-2.02) | 0.47 (0.34-0.64) | 0.66 (0.45-0.96) |
| Late (>1month) | 1.96 (1.38-2.77) | 1.7 (1.14-2.53) | 1.37 (0.87-2.16) | 1.62 (0.94-2.78) | 2.76 (2.06-3.7) | 2.61 (1.84-3.7) |
| Distance of cluster from UPHC |  |  |  |  |  |  |
| Less than 1 km |  |  |  |  |  |  |
| 1-3 km | 0.96 (0.69-1.34) | 1.31 (0.87-1.99) | 1.04 (0.69-1.58) | 1.06 (0.6-1.86) | 1.53 (1.18-1.99) | 1.7 (1.19-2.42) |
| >3 km | 1.12 (0.81-1.55) | 1.31 (0.87-1.97) | 1.44 (0.97-2.13) | 1.9 (1.13-3.19) | 1.17 (0.9-1.53) | 1.37 (0.94-2) |
| New HTN Diagnosis (vs previously known HTN) | 0.64 (0.46-0.91) | 0.55 (0.37-0.82) | 2.24 (1.54-3.27) | 2.33 (1.49-3.63) | 1.51 (1.17-1.95) | 1.03 (0.75-1.41) |
| New DM Diagnosis (vs previously known DM) | 0.55 (0.3-1.02) | 0.55 (0.29-1.06) | 0.97 (0.51-1.85) | 1.18 (0.56-2.47) | 1.2 (0.81-1.8) | 1.38 (0.87-2.19) |
| Overweight or Obese (vs Normal) based on BMI | 1.14 (0.83-1.57) | 0.91 (0.61-1.37) | 0.88 (0.59-1.31) | 1.03 (0.6-1.75) | 0.83 (0.64-1.09) | 1.02 (0.71-1.47) |
| Waist Circumference | 1.25 (0.9-1.76) | 1.74 (1.09-2.77) | 0.66 (0.45-0.98) | 0.93 (0.52-1.66) | 0.64 (0.5-0.83) | 0.97 (0.66-1.43) |

Those linked to Public health facilities (Group A) are referent; OR=Odds ratio; ASHA=Accredited social health activist; HTN=Hypertension; DM=Diabetes; BMI=Body mass index; UPHC=Urban Primary health centre.

We interviewed 167 of the randomly selected 192 individuals in detail to understand potential barriers and facilitators for non-linkage to public health facilities. The participants had overall poor knowledge about risk factors and unfavourable attitude towards availing CVD risk reduction services across all groups. Approximately three in four individual in each group were not aware about tobacco being risk factor for hypertension, while one in two individual was not aware about role of obesity in diabetes and hypertension. Non-linked individuals were in denial as they considered themselves as not having risk-factors or not identifying need for risk-factor modification. They also reported poor family and social support. Their denial about their risk-status is indirectly suggested by them not-identifying much with health-system barriers. About half of those who were not linked reported that their healthcare provider didn’t suggest any risk reduction measure or a drug therapy for them. Most individuals who were linked to private care providers identified health-system barriers with public sector, and also acknowledged that drugs in private sector are expensive. Individuals who were linked (group A and B) identified more individual level and health-system barriers. (Figure 2)

**Figure 2.**
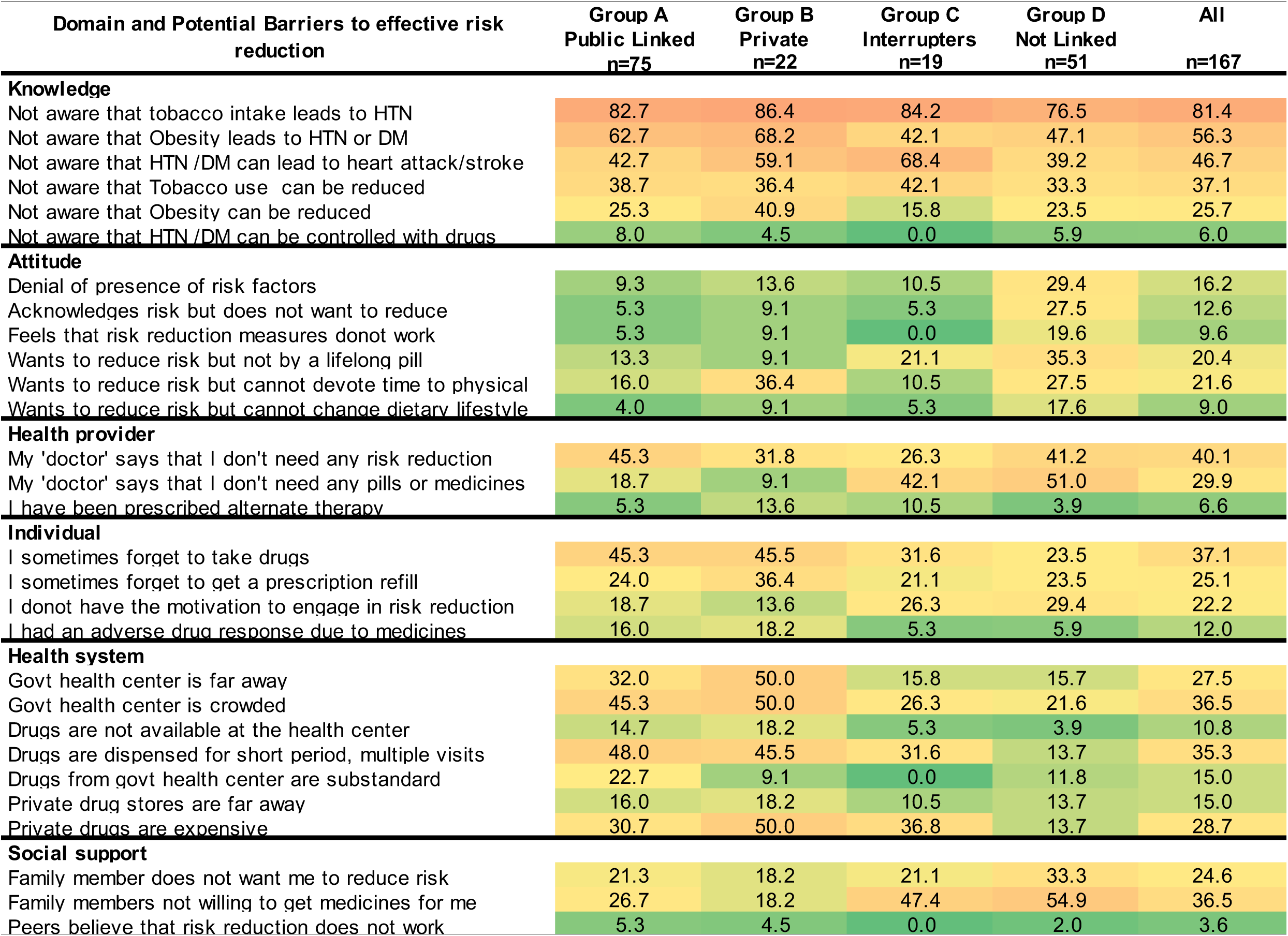
Potential barriers for linkages to public primary-care facilities. Legend : All Numbers indicate proportion of individuals in whom specified barrier is operative. This is also indicated by colour shades: Shades of Red (>60%), shades of orange (40-60%), shades of yellow (20-40%), and shades of green (0-20%). A higher proportion indicates that barrier is identified in larger number of individuals.

## Discussion

In the current study, where all individuals aged 30 years and above in the community were screened to fill the unawareness gap, six months later a little more than half of all positively screened were on pharmacotherapy from either public or private sector providers. Most of those who were on medication, were obtaining these from the public-sector. Individuals who did not get-linked to a health-care facility were more likely to be young, with poor economic status, and living farther from the UPHC. Their engagement with CHWs was also late. Those who were on pharmacotherapy identified health-system as a barrier, those not-linked identified more with poor reinforcement by family, peers, and health-care providers. Non-linked individuals were also twice more likely to deny presence of risk factor in them, or refusal to modify them, both being components of attitude domain. Modification in attitudes, social support, and provider reinforcement would be required to improve linkages.

Initiation of therapy for hypertension and diabetes is necessary to prevent CVDs. After initial screening of asymptomatic chronic diseases like hypertension or diabetes, elevated biological values need to be confirmed to ensure diagnostic certainty. Subsequently, those detected with a disease-condition need to be initiated on pharmacotherapy. Ensuring initiation of pharmacotherapy in positively screened individuals, who are otherwise asymptomatic, is challenging. We need to overcome both internal (related to awareness and acceptability) and external (related to availability and affordability of medication) barriers to ensure initiation of preventive measures.[10] Attitude of an individual reflects their preferential ways of thinking and doing in a context. Many individuals detected with a risk-factor have a ‘fear of unknown’ (in absence of objective knowledge) about NCD ‘risk’ and hence an attitudinal vulnerability may be sensed.

One decade ago, it was reported from UK that 5% of the hypertensive patients fail to initiate treatment after prescribing, and about 50% default within 1 year of treatment initiation.[16] Another study from Canada reported the detection-initiation gap in hypertension to be 18%, and discontinuation rate to be 5% in one year.[17] Medicare beneficiaries in United States were reported to have about 21% annual discontinuation rate for anti-hypertensive drugs.[18] Discontinuation rates for oral hypoglycaemic medication were reported to be as high as 49% at end of one year from another Canadian study, however re-initiation rates in the subsequent year were high.[19] In a hypertension adherence promotion trial from Nigeria, drop-out rates were reported to be about 12%, much lower than many high-income countries.[20] In our study detection-initiation gap was high (more than 30%), and discontinuation rate was modest at about 10% at the end of six months. There is a paucity of estimates from low-and-middle-income country settings about such gaps in detection to initiation and characteristics of non-initiators thereon.

In this study, those who were non-initiators were younger men compared to others. This could be explained by the unfavourable attitude towards presence of risk factors, lack of motivation to change the risk state or continue lifelong medication. It may also be related to likelihood of younger men to have occupational priorities, which makes it difficult for them to make multiple visits to the health-facility for their prescription refills. Those who are less-wealthy are also more likely to be engaged in multiple income generation activities, leading to long working hours, including weekends. Also these men got engaged late by CHWs which may have various reasons. We argue that young men may have the perception that CHWs, who are female health-volunteers, cater to health issues of “women” rather than of “men”. Moreover, CHWs usually visit households during the day-time, when most young men are not at home. Health-promotion activities in urban slums are much restricted in late-evenings both due to safety concerns, and as CHWs have to cater to their own families too. Concurrently, coverage of women has been higher in screening as well as follow-up stages in our study. Measures such as evening and holiday camps, and mop-up campaign involving male volunteers were undertaken for increasing linkage of more men to health-facilities for treatment initiation and continuation, however it had limited success. Skewed distribution of female participation was also reported in a female CHW led trial for blood pressure reduction conducted in Nepal.[21]

The variables favouring or disfavouring linkage in this study may also be thought in reference to Andersen model of total patient delay.[22] This model breaks the delay intervals into dimensional components like appraisal, illness, behavioural, and scheduling delays. Younger individuals also have a comparative shorter duration of diagnosis that may lead to ignorance and consequent treatment-neglect. Asymptomatic nature of diseases also adds to this ignorance. Fear of loss of control, impatience, and competing priorities also force them to develop a selective blindness to self. Early ASHA engagement may give the positive signal to participants about the importance of early diagnosis and treatment and an activist attitude of health system towards the ailment and vice versa. This in turn may prompt the individual to transform from ‘slack deterrent individual’ to ‘tense motivated individual’ who wants to actively seek treatment.

In comparison to those who seek care from public sector, individuals under treatment from private care facilities were more likely to be men, belonging to higher wealth quintile and previously known to have hypertension. While knowledge, attitude, and individual level barriers in both these subgroups were comparable, those who sought treatment from private sector identified with greater reinforcement from health-care providers, and family members. Interestingly, privately-linked individuals identified distance and overcrowding in the health-facilities as a greater barrier, rather than questioning the availability or quality of medicines available from public-sector facilities.

Prevention of CVD requires that the public health facilities should have at least three anti-hypertensive drugs (Angiotensin-receptor blocker or an angiotensin converting enzyme inhibitor, Calcium channel inhibitor, and a thiazide diuretic), two oral hypoglycemics (A biaguanide and a sulphonylurea), a statin and low dose aspirin. These have also been identified by WHO-PEN as essential ingredients for NCD care.[14] The WHO Package of Essential Noncommunicable Disease Interventions (WHO PEN) for primary care in low-resource settings describes set of cost-effective interventions and resources for NCDs which can be used even in resource poor settings. Previous studies have demonstrated that availability of these medications in private pharmacies in India is comparable to high-income-countries, however households are unlikely to afford these due to lower paying capacity.[6] In our study 17.3% of high risk individuals preferred private sector for their drug prescription, and understandably they were in higher wealth quintiles as compared to those linked to public health facilities.

Various factors influence acceptance and persistence with life-long pharmacotherapy, and its patterns are heterogenous.[23] Detection-to-initiation time interval for anti-hypertensive drug therapy is longer in younger, as compared to older hypertensive individuals.[24] Poor economic status, and poor disease control are strong predictors of discontinuation of pharmacotherapy.[25] Regular medication use in chronic diseases requires a daily, lifelong, repetitive, habit-forming behavior. Behavioral theories suggest that individuals with medication taking habit strength are most likely to exhibit long term adherence.[26] Hence individuals who take their medications regularly when initiated on drug therapy, are most likely to be adherent to their medication over a long-term.[27] To ensure a perpetual habit-forming behavior, a strong early reinforcement needs to be advocated for chronic pharmacotherapy.

We need robust mechanisms to monitor adherence especially when a large number of individuals are likely to be screened and treated in public sector.[28] We also need an efficient health-system that ensures continual access to medication, with minimum disruption of occupational priorities. Recent guidelines for hypertension, and higher CVD risk in South-Asians, advocates a more aggressive management of hypertension and diabetes mellitus.[29] Various systematic reviews have recorded numerous successful interventions, to overcome barriers at individual, family, community, provider, and health-system levels.[30] Interventions that addressed barriers at multiple-levels were more successful than the interventions that focused on a single or fewer barriers.[31] Some of the cost-effective solutions could include improved information and behavior change measures by community health workers, reinforcements by family and providers, improved drug packaging, accessibility, and monitoring mechanisms at the health-system levels.

### Strengths and Limitations

A key strength of the study is its implementation through stakeholders of existing public health system who are expected to perform these activities under NPCDCS. This study was limited to urban slum clusters from a single city, however we believe that health-infrastructure is broadly similar in such settings elsewhere. All CHWs were newly trained in non-communicable disease work, while their primary training is in reproductive and child health service delivery. This new area of work for them was also a competing priority relative to their previous routine tasks in reproductive and child health domains. This limits focus of CHWs on non-communicable disease related work, but is also a reflection of “real-world” situation in various developing countries.

## Conclusions

CHW led strategy for screening, treatment linkage and follow up of hypertension and diabetes for CVD reduction are feasible. However, a large gap exists between high-risk condition detection to treatment-initiation. This subgroup of newly detected high risk group would need distinct efforts directed to address their state of denial. Also, strategy to decrease treatment initiation to discontinuation trajectory needs to be developed. This can be achieved through improved information and behavior change measures by community health workers, reinforcements by family and providers, improved accessibility, continued drug supply, newer ways of drug packaging and monitoring mechanisms at the health-system levels.

## Supporting information

STROBE Checklist

## Data Availability

Raw data of this study is not deposited in any public repository. However, anonymized raw data of this study would be available to academicians or researchers on request to corresponding author.

## Ethical approval

The study design was approved by the Institutional Human Ethics Committee of All India Institute of Medical Sciences, Bhopal(Ref: IHEC-LOP/2017/EF00045)

## Informed consent

Participant Information Sheet in Hindi language was provided to each participant. All participants provided written informed consent prior to initiation of any study procedures.

## Author contributions

RJ conceived the study; RJ, APP and SK developed the protocol; RA, KD, IT, VM and NS acquired data, AJ, SA, SK, APP and RJ supervised data acquisition and verified diagnosis, APP, AJ and RJ analysed data and wrote first draft. All authors critically reviewed the first draft and provided inputs for its revisions.

